# Griffin: Framework for clinical cancer subtyping from nucleosome profiling of cell-free DNA

**DOI:** 10.1101/2021.08.31.21262867

**Authors:** Anna-Lisa Doebley, Minjeong Ko, Hanna Liao, A. Eden Cruikshank, Caroline Kikawa, Katheryn Santos, Joseph Hiatt, Robert D. Patton, Navonil De Sarkar, Anna C.H. Hoge, Katharine Chen, Zachary T. Weber, Mohamed Adil, Jonathan Reichel, Paz Polak, Viktor A. Adalsteinsson, Peter S. Nelson, Heather A. Parsons, Daniel G. Stover, David MacPherson, Gavin Ha

## Abstract

Cell-free DNA (cfDNA) has the potential to inform tumor subtype classification and help guide clinical precision oncology. Here we developed Griffin, a new method for profiling nucleosome protection and accessibility from cfDNA to study the phenotype of tumors using as low as 0.1x coverage whole genome sequencing (WGS) data. Griffin employs a novel GC correction procedure tailored to variable cfDNA fragment sizes, which improves the prediction of chromatin accessibility. Griffin achieved excellent performance for detecting tumor cfDNA in early-stage cancer patients (AUC=0.96). Next, we applied Griffin for the first demonstration of estrogen receptor (ER) subtyping in metastatic breast cancer from cfDNA. We analyzed 254 samples from 139 patients and predicted ER subtype with high performance (AUC=0.89), leading to insights about tumor heterogeneity. In summary, Griffin is a framework for accurate clinical subtyping and can be generalizable to other cancer types for precision oncology applications.

## Introduction

Accurate cancer diagnosis and subtype classification are critical for guiding clinical care and precision oncology. Current approaches to determine tumor subtype require a tissue biopsy, which is often difficult to obtain from patients with metastatic cancer. Therefore, at the time of recurrence or metastatic cancer diagnosis, treatment options may often be informed by clinical diagnostics from the primary tumor. However, molecular changes in the tumor can emerge during metastatic progression and in the context of therapeutic resistance. Moreover, surveying molecular changes is challenging because repeated biopsies are problematic and not routine in clinical practice for solid tumors.

Cell-free DNA (cfDNA) is DNA released into circulation by cells during apoptosis and necrosis.^1^ In patients with cancer, a portion of this cfDNA is released from tumor cells, called circulating tumor DNA (ctDNA). The analysis of ctDNA can address the challenges in tissue accessibility and has demonstrated great potential for clinical utility.^2–9^ Much of the current research and clinical efforts have focused on the detection of genetic alterations in ctDNA. Shallow coverage sequencing of cfDNA, including ultra-low pass whole genome sequencing (ULP-WGS, 0.1x), provides a cost-effective and scalable solution for estimating the tumor fraction (fraction of the cfDNA that is tumor derived) from the analysis of genomic copy number alterations.^10–13^ Sequencing analysis of genomic alterations from ctDNA have helped to distinguish molecular subsets of tumors.^14,15^ However, these genomic alterations, including somatic mutations, may not always fully explain treatment failure or identify therapeutic targets, exemplifying a major limitation of cancer precision medicine.

Tumor subtypes are often characterized by distinct transcriptional regulation, which can change during treatment resistance, leading to different clinical tumor phenotypes. For example, prostate and lung cancers may undergo trans-differentiation from adenocarcinoma to small-cell neuroendocrine phenotypes.^16–20^ For metastatic breast cancer (MBC), treatment is guided based on clinical subtypes determined by the expression of the estrogen receptor (ER), progesterone receptor (PR), and human epidermal growth factor receptor 2 (HER2), often in the primary tumor^21^; endocrine therapies are prescribed to patients with ER-positive (ER+) or PR-positive (PR+) carcinomas while patients with HER2 positive tumors are prescribed anti-HER2 drugs. Patients with tumors absent for expression of all three receptors have triple negative breast cancer (TNBC) and receive chemotherapy.^22^ However, receptor conversions during primary and metastatic disease progression have been frequently observed, including ∼20% of patient tumors switching from ER+ to ER-negative (ER-) subtypes.^23–28^ Furthermore, similar to the presence of intra-tumor genomic heterogeneity in breast cancer, mixtures of clinical subtypes may also co-exist across or within metastatic lesions in the same patient, presenting major clinical challenges.^29,30^ Therefore, accurate subtype classification and identification of transcriptional patterns underlying emergent clinical phenotype during therapy has critical implications for studying mechanisms of resistance and informing treatment decisions.

Recent studies have shown that the computational analysis of cfDNA fragmentation patterns from genome sequencing data can reveal the occupancy of nucleosomes in cells-of-origin.^31–36^ When DNA is released into the peripheral blood following cell death, they are protected from degradation by nucleosomes.^1^ At accessible genomic locations, such as at actively bound transcription factor binding sites (TFBSs) and open chromatin regions, nucleosomes are positioned in an organized manner that allows access for DNA binding proteins^37^ (Fig. 1a). This nucleosome organization results in a loss of sequencing coverage, reflecting DNA degradation at the unprotected binding site with peaks of coverage at the surrounding protected locations.

**Fig. 1.**
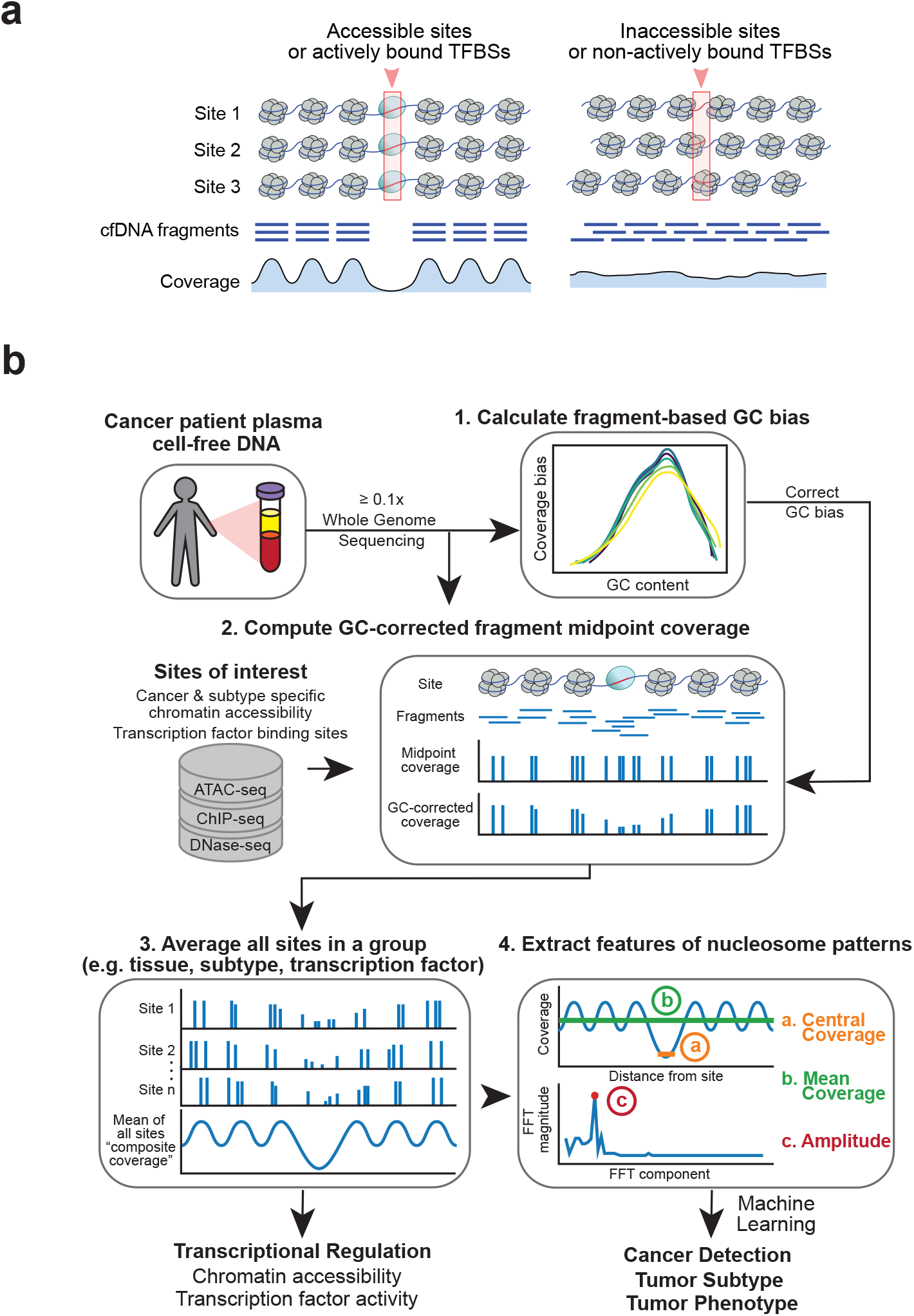
Griffin framework for cfDNA nucleosome profiling to predict cancer subtypes and tumor phenotype. **(a)** Illustration of a group of accessible sites (left panel) and inaccessible sites (right panel), such as a TFBS. The nucleosomes (in grey) are positioned in an organized manner around the accessible sites (red box; left panel), but not around the inaccessible ones (right panel). These nucleosomes protect the DNA from degradation when it is released into peripheral blood. The protected fragments from the plasma are sequenced and aligned, leading to a coverage profile which reflects the nucleosome protection in the cells of origin. **(b)** Griffin workflow for cfDNA nucleosome profiling analysis. cfDNA whole genome sequencing (WGS) data with ≥ 0.1x coverage is aligned to hg38 genome build. (1) For each sample, fragment-based GC bias is computed for each fragment size. (2) Sites of interest are selected from any assay. Paired-end reads aligned to each site are collected, fragment midpoint coverage is counted, and corrected for GC bias to produce a coverage profile. (3) Coverage profiles from all sites in a group (e.g., open chromatin for tumor subtype) are averaged to produce a composite coverage profile. Composite profiles are normalized using the surrounding region (−5 kb to +5 kb). (4) Three features are extracted from the composite coverage profile: central coverage (coverage from -30 bp to +30 bp from the site; orange ‘a’), mean coverage (between -1 kb to +1 kb; green ‘b’), and amplitude calculated using a Fast-Fourier Transform (FFT) (red ‘c’).

Applications of nucleosome profiling from cfDNA have been demonstrated for cancer detection and tumor tissue-of-origin prediction, including the analysis of shorter cfDNA fragments which tend to be enriched from tumor cells.^38–41^ While tumor subtyping from cfDNA has been explored in prostate cancer by analyzing TFBS locations^42^, to our knowledge there have not been demonstrations of subtype classification from cfDNA in other cancers. Specifically, predicting histological subtypes in breast cancer has not been shown from cfDNA. Furthermore, current cfDNA nucleosome profiling approaches have not been optimized for ULP-WGS data. Studying the clinical phenotype of tumors from ctDNA remains challenging due to lack of robust computational methods but has obvious potential clinical benefits for guiding treatment decisions in patients with metastatic cancer.

In this present study, we developed a computational framework called Griffin to classify tumor subtypes from nucleosome profiling of cfDNA. Griffin overcomes current analytical challenges to profiles the nucleosome accessibility and transcriptional regulation from the analysis of standard cfDNA genome sequencing, including ULP-WGS (0.1x) coverage. Griffin employs a novel GC correction procedure that is specific for DNA fragment sizes and therefore unique for cfDNA sequencing data. We applied Griffin to perform cancer detection and tumor tissue-of-origin analysis with high performance. Then, we demonstrate the first application of breast cancer ER subtyping from cfDNA, showing strong classification accuracy and insights into tumor heterogeneity and prognosis, all achieved from analysis of ULP-WGS data. Overall, Griffin is a generalizable framework that can detect molecular changes in transcriptional regulation and chromatin accessibility from cfDNA and possibly direct personalized treatment to improve patient outcomes.

## Results

### Griffin framework for nucleosome profiling to predict tumor phenotype

We developed Griffin as an analysis framework with a new GC correction procedure to accurately profile nucleosome occupancy from cfDNA. Griffin processes fragment coverage to distinguish accessible and inaccessible features of nucleosome protection (Fig. 1a). Griffin is designed to be applied to whole genome sequencing (WGS) data of cfDNA from patients with cancer to quantify nucleosome protection around sites of interest and is optimized to work for ULP-WGS data (Fig. 1b). Sites of interest can be selected from various chromatin-based assays, such as from assay for transposase-accessible chromatin using sequencing (ATAC-seq) and are tailored to address specific problems including cancer detection and tumor subtyping.

The analysis workflow begins with computing the genome-wide fragment-based GC bias for each sample. Then, for the region at each site of interest, the fragment midpoint coverage is computed and reweighted to remove GC biases (Methods). Midpoint coverage rather than full fragment coverage is used because it produces higher amplitude nucleosome protection signals (Supplementary Fig. 1). Next, a composite coverage profile is computed as the mean of the GC-corrected coverage across the set of sites specific for a tissue type, tumor type, transcription factor (TF), or any phenotypic comparison of interest. By examining these coverage profiles around known cancer-specific and blood-specific TFs, we identified three quantitative features that distinguish a site as accessible and inaccessible: (a) the coverage in the window between -/+ 30 bp (‘central coverage’), where lower values represent increased accessibility, (b) the coverage in a window between -/+ 1000 bp (‘mean coverage’), and (c) the overall nucleosome peak amplitude calculated using Fast Fourier transform (FFT, ‘amplitude’). These features can be used to quantify transcription factor activity or chromatin accessibility and be used as features for detection of cancer, tumor subtyping, or studying other phenotypes of interest.

### Griffin reduces GC biases enabling detection of tissue specific accessibility

A novel aspect of Griffin is the implementation of a fragment-based GC bias correction. At open chromatin regions, especially at TFBS, GC-content is non-uniform, which leads to GC-related coverage biases (Fig. 2a).^43^ GC bias varies between samples and between different fragment lengths within a sample^44^ (Fig. 2b), which can have a major impact on nucleosome accessibility prediction (Fig. 2c). To correct for this GC bias, for each sample and each fragment length, Griffin computes the global estimated mean fragment coverage (“expected”) using a fragment length position model^44^ (Methods, Fig. 2b). Then, when calculating coverage around sites of interest, each fragment is assigned a weight based on the global expected coverage. This correction eliminates unexpected increases (or decreases) in coverage at binding sites, removing technical biases to enhance the epithelial tissue-associated accessibility signals when analyzing WGS (9-25x, Fig. 2c) cancer patient cfDNA and ULP-WGS (0.1-0.3x, Fig. 2d).

**Fig. 2.**
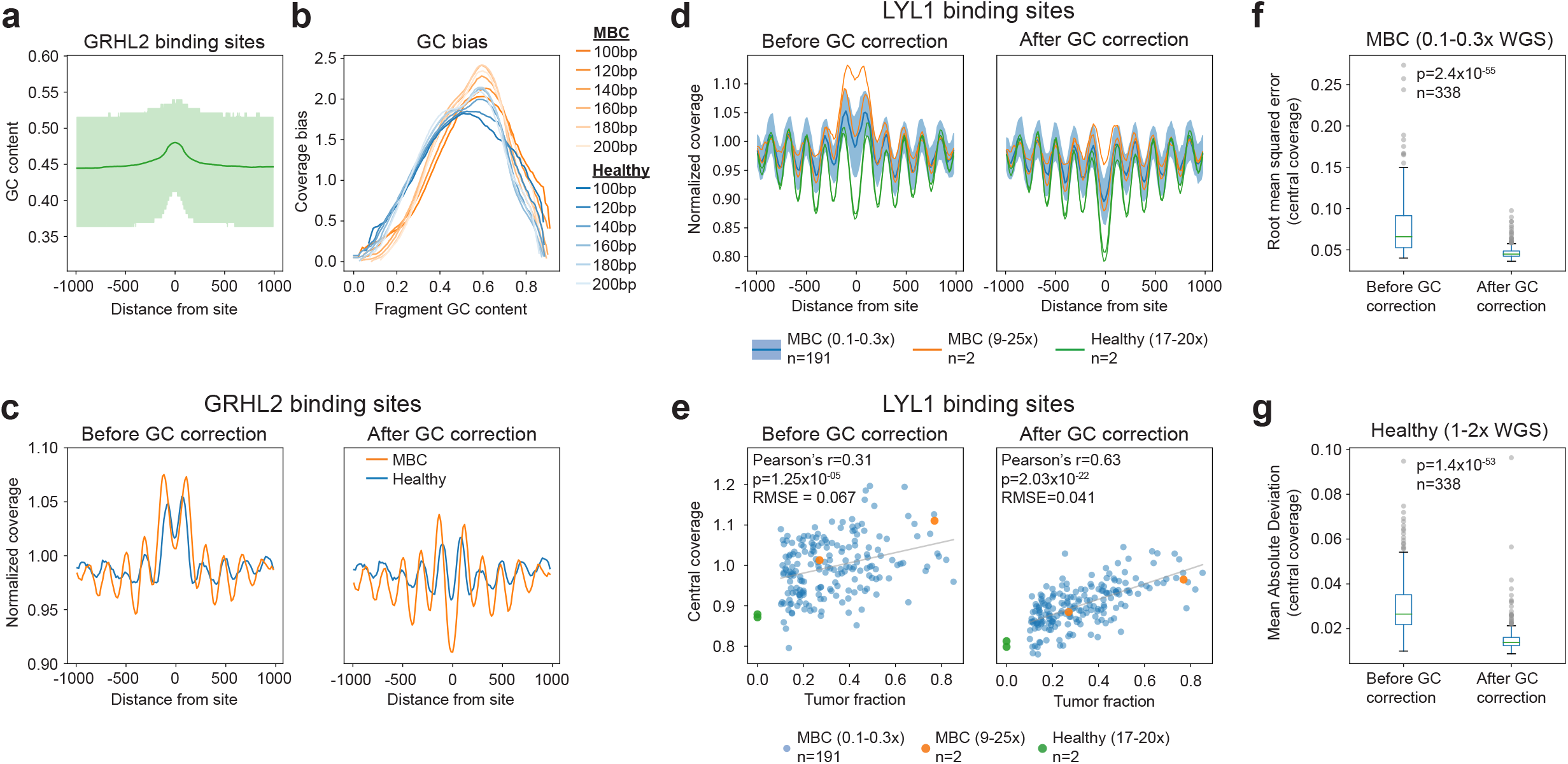
Griffin GC bias correction improves detection of tissue specific accessibility from cfDNA. **(a)** Aggregated GC content at 10,000 GRHL2 binding sites and its surrounding 2kb region. Mean GC content (line) and interquartile range (green shading) are shown. **(b)** cfDNA GC bias is unique to each sample and each fragment length. GC bias computed for cfDNA from a healthy donor (HD_46; blue shades) and a metastatic breast cancer (MBC_315; orange shades) sample are shown for various fragment sizes. **(c)** Composite coverage profile of 10,000 GRHL2 binding sites before and after GC correction, shown for HD_46 (blue) and MBC_315 (orange). Before GC correction, the ‘central coverage’ has a higher value due to effects of GC bias which can obscure differential signals between samples. After GC correction, the central coverage of the MBC sample has lower value, which is consistent with increased GRHL2 activity in breast cancer but not immune cells making up the healthy donor sample. **(d)** Composite coverage profiles of 10,000 LYL1 sites before and after GC correction, shown for two MBC samples with deep WGS (9-25x, orange), two healthy donors (17-20x, green), and 191 MBC samples with ULP-WGS (0.1-0.3x, blue). Median +/- IQR of 191 ULP-WGS samples is shown with blue shading. Lower ‘central coverage’ corresponding to greater site accessibility in the healthy donor samples is expected because LYL1 is a transcription factor associated with hematopoiesis. **(e)** cfDNA tumor fraction and central coverage correlation for LYL1, shown for ULP-WGS (0.1-0.3x, n=191) and WGS (9-25x, n=2) of MBC and healthy donors (17-20x, n=2) samples. cfDNA contains a mixture of tumor and blood cells; therefore, central coverage value is expected to be positively correlated with tumor fraction (lower represents increased accessibility). After GC correction, the correlation (for the MBC ULP-WGS samples) is much stronger based on Pearson’s r correlation coefficient. Root mean squared error (RMSE) of the linear fit is shown. **(f)** Boxplots showing the distribution of the RMSE (linear fit between central coverage and tumor fraction in the MBC ULP-WGS dataset [0.1-0.3x, n=191]) across the 338 TFs, before and after GC correction. The boxed range represents the median ± IQR, whiskers represent the range of the non-outlier data (maximum extent is 1.5x the IQR). Outliers are plotted in grey. p-value was calculated using the Wilcoxon signed-rank test (two-sided). **(g)** Boxplots showing the distribution of the mean absolute deviation (of the central coverage across 215 healthy donors [1-2x WGS]) across the 338 TFs, before and after GC correction. Box elements are the same as (f). p-value was calculated using the Wilcoxon signed-rank test (two-sided).

To test the performance of nucleosome profiling following Griffin GC-bias correction, we compared the estimated TFBS accessibility with the amount of tumor-derived DNA (i.e. tumor fraction) predicted by ichorCNA for ULP-WGS data from 191 MBC cfDNA samples with ≥ 0.1 tumor fraction.^10^ We expect the tumor fraction to be negatively corrected with the central coverage around tumor-specific sites, and positively correlated for blood-specific sites. For a blood specific TF, LYL1, we observed that the central coverage at TFBSs was positively correlated with tumor fraction before GC correction (Pearson’s r=0.31) as expected, but this correlation was much stronger after GC correction (Pearson’s r=0.63, Fig. 2e). For a tumor-specific TF, GRHL2, we observed a negative correlation between the central coverage and tumor fraction, as expected (Pearson’s r=-0.63, Supplementary Fig. 2). The mean coverage and amplitude features are also correlated to tumor fraction but appeared to be less influenced by GC bias (Supplementary Fig. 2, Supplementary Data 1). Similar correlations between nucleosome profile features and tumor fraction following GC correction were also observed for blood and cancer specific DNase I hypersensitivity sites (DHSs) (Supplementary Fig. 2).

To quantify how GC correction reduces signal variability between samples, we examined the central coverage in the 191 MBC cfDNA ULP-WGS samples for 338 TFs in the Gene Transcription Regulation Database (GTRD).^42,45^ For each factor, we compared the variability between the central coverage and tumor fraction using the root mean squared error (RMSE) from a linear regression fit before and after GC correction. For LYL1, the RMSE decreased (0.067 to 0.041), indicating less inter-sample variation in the data after GC correction (Fig. 2e). Similarly, for 325 (96.1%) TFs, the RMSE was decreased after GC correction, indicating reduced inter-sample variability after accounting for the correlation between tumor fraction and central coverage (two-sided Wilcoxon signed rank test p = 2.4×10^−55^, test statistic = 472, Fig. 2f, Supplementary Data 1). Additionally, we examined the central coverage for the 338 TFs in a cohort of 215 healthy donors^38^ before and after GC correction. Because healthy donor samples have no tumor content, we evaluated the mean absolute deviation (MAD) for each TF to compare inter-sample variability. We found that the MAD decreased after GC correction for 324 (95.8%) TFs (two-sided Wilcoxon signed rank test p = 1.4×10^−53^, test-statistic = 940, Fig. 2g, Supplementary Data 2), indicating lower inter-sample variability for nearly all TFs. Altogether, these results suggest that the novel GC correction in the Griffin framework reduces the variability in chromatin accessibility signals due to GC biases between samples and allows for improved detection of tissue specific accessibility in ULP-WGS data.

### Griffin analysis at TFBS enables accurate cancer detection and tissue-of-origin prediction

To determine if Griffin can perform cancer detection, we analyzed a published WGS (1-2X) dataset of cfDNA samples from healthy donors (n = 215) and cancer patients (n = 208).^38^ We generated nucleosome profiles around TFBSs for the 338 TFs using nucleosome sized (100-200bp) fragments and extracted three features from each profile (central coverage, mean coverage, and amplitude) for a total of 1014 features. Using logistic regression, we achieved a high performance for predicting the presence of cancer with an area under the receiver operating curve (AUC) of 0.96 (Fig. 3a, Supplementary Data 3). We achieved the highest prediction performance for lung and ovarian cancers (AUC=1.00) and the lowest for pancreatic cancer (AUC=0.90). We also observed high performance for stage IV cancers (AUC=0.99) but maintained great performance for stage I cancers (AUC=0.94, Fig. Supplementary Fig. 3). The performance was likely reflective of the higher tumor fractions observed in late-stage cancer relative to early-stage cancer. We observed higher performance for samples with tumor fraction ≥ 0.05 (AUC 1.0) than samples with undetectable tumor (0 tumor fraction, AUC=0.94, Supplementary Fig. 3). We also observed similar performance with Griffin analysis around DNase I Hypersensitivity Sites (DHS) (AUC=0.91, Supplementary Fig. 3).

**Fig. 3.**
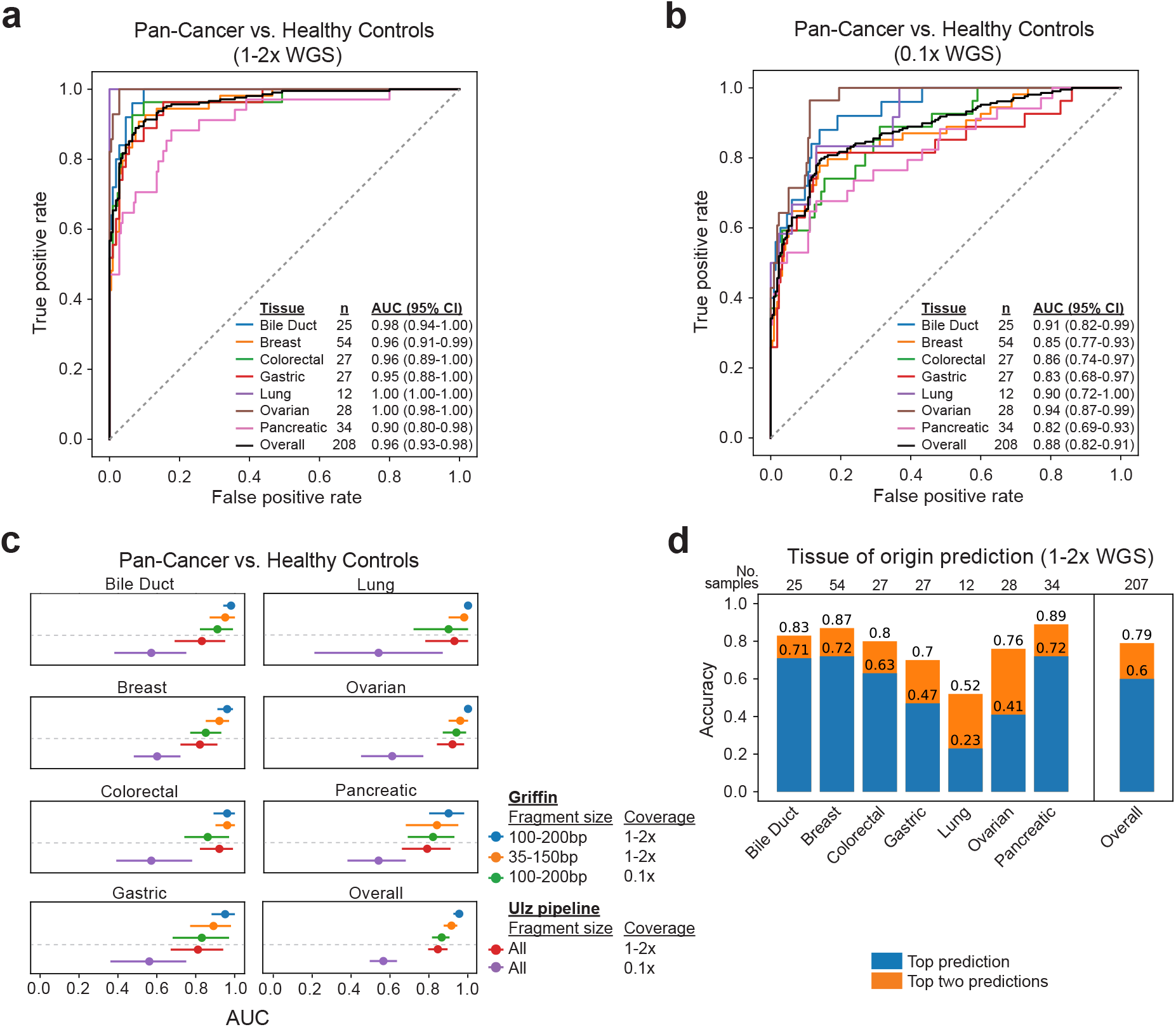
Griffin enables accurate cancer detection and tissue-of-origin prediction. **(a)** Receiver operator characteristic (ROC) curve for logistic regression classification of cancer (n=208) vs. healthy controls (n=215)^38^ using nucleosome profiles around TFBSs in 1-2x WGS data. ROC for each cancer type vs. healthy are shown. 95% confidence intervals (CIs) were obtained by bootstrapping. Duodenal cancer (n=1) is not shown. **(b)** ROC for logistic regression classification of cancer using the same TFBSs feature set applied to the same dataset downsampled to 0.1x WGS coverage. **(c)** Area under the ROC curve (AUC) values for logistic regression models using different feature sets collected from nucleosome profiling around TFBSs. The fragment size range, sample coverage, and nucleosome profiling tool (Griffin and Ulz pipelines) are indicated. 95% CIs were obtained by bootstrapping. **(d)** Accuracy of a multinomial logistic regression model used to predict tissue-of-origin in 207 cancer patients (duodenal cancer was excluded). The accuracy of the top prediction and top two predictions by the model are shown for each individual cancer type and overall, for all cancer types combined.

To test the ability to detect cancer at ULP-WGS coverage (0.1x), we applied Griffin to the same cfDNA data downsampled to 0.1x coverage and achieved a performance with AUC of 0.88 (Fig. 3b). Next, because fragments <150bp are enriched for tumor derived DNA^38^, we tested whether using only shorter fragments might improve our ability to detect cancer in this framework, we applied Griffin to analyze only 35-150bp fragments at the same TFBSs and observed a decreased performance (AUC=0.93, Supplementary Fig. 3). Finally, we compared our results with the method by Ulz et al.^42^, which analyzed cfDNA fragments of all lengths at TFBSs. Across all cancer types, Griffin using nucleosome-sized or short fragments and ULP-WGS coverage had higher detection performance (Fig. 3c, Supplementary Fig. 3). This demonstrates that Griffin can detect cancer accurately using various sites from chromatin-based assays and cost-effective ULP-WGS of cfDNA.

Next, we tested the ability of Griffin to predict the cancer tissue of origin from cfDNA. Using Griffin nucleosome profile features around the TFBSs for the 338 TFs, we applied a multinomial logistic regression to predict the cancer type of each sample. The top prediction was correct for 60% of samples. When the top two predictions were considered, 79% of the samples were correctly classified (Fig. 3d). Overall, we show that Griffin can be used for highly accurate cancer detection from cfDNA even when using ULP-WGS coverage and that Griffin can be used for tissue of origin prediction.

### Griffin enables accurate prediction of breast cancer subtypes from ultra-low pass WGS

Breast cancer tumor classification relies on accurate clinical determination of hormone receptor status primarily by immunohistochemistry (IHC) to quantify the expression of ER, but no ctDNA approach exists for this application. We set out to determine whether Griffin can be used to predict ER subtype status from ULP-WGS (0.1x) of cfDNA from MBC patients. We analyzed 254 samples^10,11^ with tumor fraction greater than 0.05 from 139 patients. First, we inspected the Griffin profiles at TFBSs for key factors, including ESR1, FOXA1, and GATA3, which are known to be associated with ER positive tumors.^46^ We observed that these TFBSs were more accessible in cfDNA samples from patients with ER+ metastases compared to ER-; central coverage was negatively correlated with tumor fraction for ER+ samples only (Pearson’s r < -0.35, p < 4.2×10^−4^, Supplementary Fig. 4). To predict ER status, we initially built a logistic regression classifier using features from the Griffin profiles for all 338 TFs and achieved an accuracy of 0.68 (AUC of 0.74, Supplementary Fig. 5). We also used TFBSs features computed by the Ulz method for ER subtyping and observed an accuracy of 0.55 (AUC=0.58, Supplementary Fig. 5), likely because it was not designed for ULP-WGS data.

Next, we used a more tailored site selection approach by analyzing regions of differential chromatin accessibility. Using ATAC-seq data generated from 44 ER+ and 15 ER-primary breast tumors by The Cancer Genome Atlas (TCGA)^47^, we identified open chromatin sites that were specific to each ER subtype (Methods, Fig. 4a, Supplementary Data 4). ER+ specific sites (n=27,359) were enriched for the TFBSs of ESR1, PGR, FOXA1 and GATA3, and ER-specific sites (n=24,861) were enriched for the TFBSs of STAT3 and NFKB1 (Supplementary Data 5). We observed differences in coverage profiles between ER subtype-specific sites that were shared and not shared with accessible chromatin in hematopoietic cells^48^ and analyzed them separately (Fig. 4b, Supplementary Fig. 6).

**Fig. 4.**
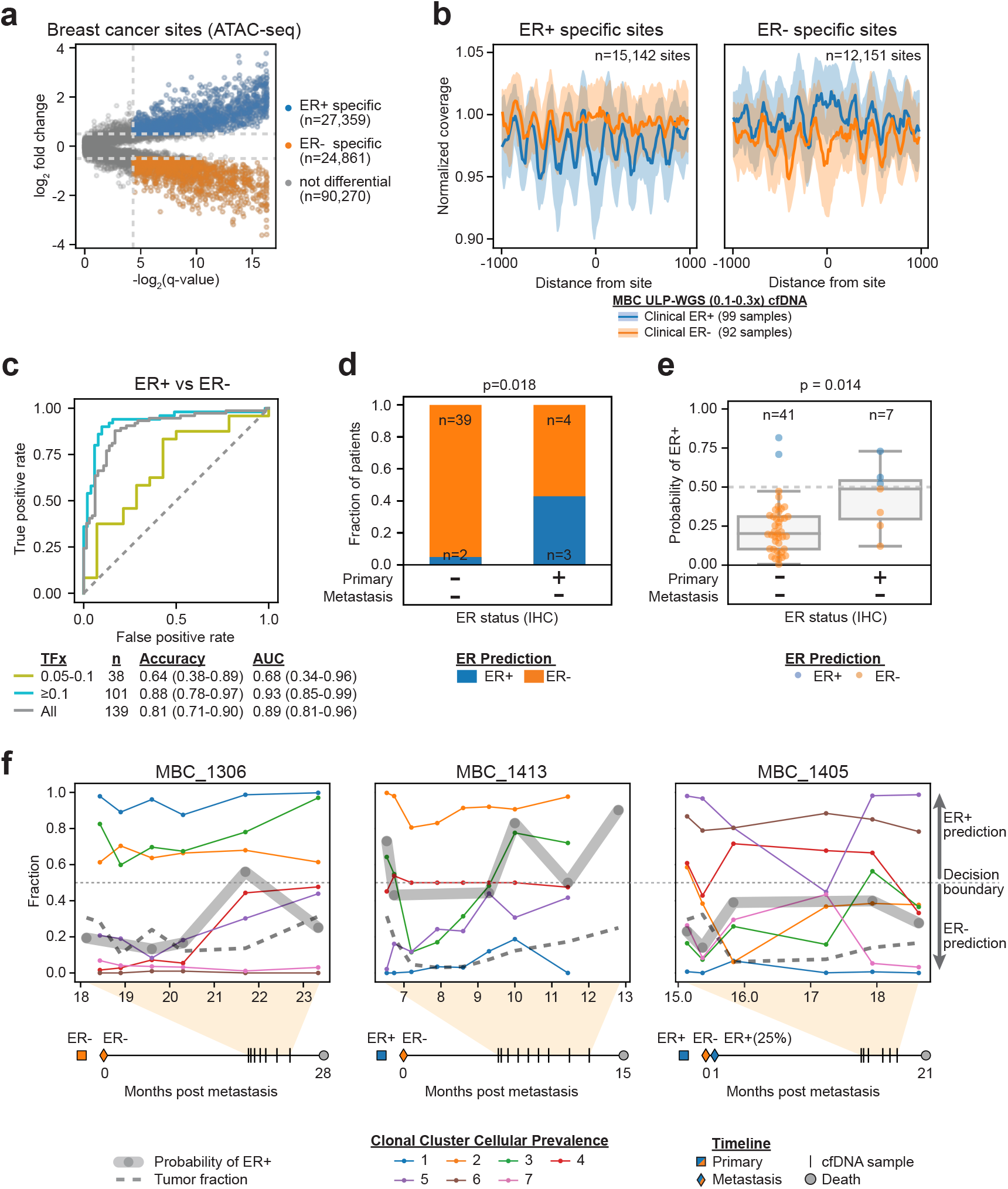
Griffin enables accurate prediction of breast cancer estrogen receptor subtypes from ultra-low pass WGS. **(a)** ER+ and ER- specific open chromatin sites were selected from assay for transposase-accessible chromatin using sequencing (ATAC-seq) data from ER+ (n=44) and ER-(n=15) breast tumors in The Cancer Genome Atlas (TCGA).^47^ Sites were selected using a Mann-Whitney-U (two-sided) test with Benjamini-Hochberg p-value adjustment (q-value) for each site, and the log2 fold change was also calculated. Sites with a q-value <0.05 and a log2 fold change of >0.5 or <-0.5 were considered differential. **(b)** Composite coverage profiles (median ± IQR) for ER+ specific (n=15,142) and ER- specific (n=12,151) sites are shown for MBC patients (≥ 0.1 tumor fraction) separated by clinical ER status (ER+, n=99; ER-, n=92). Sites shared with hematopoietic cells were excluded.^48^ **(c)** Receiver operator characteristic (ROC) curve for a logistic regression model predicting ER+ and ER- subtype. ROC curve, accuracy and AUC are shown for all patients and for patients grouped by tumor fraction (TFx), 0.05-0.1 and ≥0.1. 95% CIs were obtained by bootstrapping. For patients with multiple samples, the first sample with tumor fraction >0.05 was used. **(d)** Subtype prediction in patients with metastatic ER- breast cancer separated by clinical primary tumor ER status. P-value was calculated using a Fisher’s exact test (two-sided). **(e)** Boxplot showing the distribution of probabilities of ER+ for the same patients as in (d). The boxed range represents the median ± IQR, whiskers represent the range of the non-outlier data (maximum extent is 1.5x the IQR). All individual points are plotted. P-value calculated using ANCOVA with tumor fraction as a covariate. **(f)** Cellular prevalence of clonal clusters, ER+ prediction probability (grey line), and tumor fraction (dashed line) for multiple plasma samples shown for patients, MBC 1306, MBC 1413, and MBC 1405. Cellular prevalence was obtained from a previous study using PyClone analysis of whole exome and targeted panel sequencing of the same samples; analysis was performed independently for each patient.^49^ Decision boundary for ER+ (≥0.5) and ER- (<0.5) is indicated with dotted line. Timelines in months from metastatic diagnosis to death are shown for each patient. For patient MBC_1405, two metastatic biopsies were taken shortly after metastatic diagnosis. One was ER- (Chest wall lesion, biopsy taken at metastatic diagnosis), and one was moderately ER+ (25% ER staining, bone lesion, taken 26 days after diagnosis). This patent was considered ER+ for the purpose of the classifier (see Methods) but predicted as ER- for all timepoints.

We applied Griffin to profile nucleosome accessibility at these four sets of ER subtype-specific accessible chromatin sites, extracting a total of 12 features (Fig. 4b, Supplementary Fig. 6). We built a logistic regression classifier to predict ER subtype from these chromatin accessibility features and achieved an overall accuracy of 0.81 (AUC=0.89, n=139) (Methods, Fig. 4c). The performance was higher for samples with high tumor fraction (accuracy 0.88, AUC=0.93, n=101, tumor fraction ≥ 0.1) compared to those with lower tumor fraction (accuracy 0.64, AUC=0.68, n=38, tumor fraction 0.05 to 0.1) (Fig. 4c). Repeating the analysis using only short fragments (35-150bp) did not improve the performance (accuracy 0.66, AUC=0.71), likely due to further reduced fragment coverage (Supplementary Fig. 5). These results illustrate the utility of using chromatin accessibility for cancer subtyping from ULP-WGS data and showcase the first application of ER status prediction in breast cancer from cfDNA.

### Analysis of ER status from cfDNA reveals tumor subtype heterogeneity

To further investigate the ER predictions, we inspected the classification results for 48 of the patients with an ER-metastasis, known primary ER status, and a tumor fraction of ≥0.1. In 41 patients with where the primary and metastasis were both ER-by IHC, we predicted 39 (95.1%) patients to have ER-subtype. Intriguingly, in the seven patients who had clinical primary ER+ and metastatic ER-status (i.e., ER loss), three (42.9%) were predicted to be ER+, and this higher prevalence of ER+ prediction for this patient group was statistically significant (two-sided Fisher’s exact test p = 0.018, Fig. 4d). We also observed that the predicted probability of ER+ was higher in the patients with ER loss than the patients with ER-primary and metastasis, and this was statistically significant even after accounting for tumor fraction (analysis of covariance, p=0.014). These results suggest that there may be residual ER+ tumor features in the ER loss patients or that Griffin analysis may be capturing a heterogeneous mixture of ER subtypes from ctDNA.

To further assess whether this observation may be due to tumor heterogeneity, we examined ULP-WGS samples from six TNBC patients receiving treatment with Cabozantinib who had plasma collected at different timepoints and had clonal dynamics analysis performed previously using subclonal somatic mutations from ctDNA.^11,49^ Overall for all six patients, the ER+ probability followed closely to the trends of multiple clones over time (Fig. 4f, Supplementary Fig. 7). In patient MBC_1306, ER+ probability tracked closely with the clonality of clonal cluster 4, as estimated by the cellular prevalence^50^, particularly at 21.7 months post-metastasis where both increased (Fig. 4f). Two of these six patients (MBC_1413 and MBC_1405) had known ER loss for at least one metastasis. Interestingly in both cases, the ER+ probability fluctuated over time, but tracked with one or more of the genomic clones (Fig. 4f). In patient MBC_1413, who had an ER+ primary and ER-metastasis, we noted the ER+ probability tracked closely with the cellular prevalence of clonal cluster 3, including the coincident 0.4 ER+ probability increase with a 30% (cluster 3) expansion at 10 months post-metastasis (Fig. 4g). Patient MBC_1405 had an ER+ primary and both ER- and ER+ metastatic biopsies but was considered ER+ status despite having only 25% expression by IHC. While all five timepoints from this patient were predicted to be ER-, the ER+ probability tracked with both clonal clusters 3 and 4. Furthermore, the proximity of the predicted ER+ probabilities near the decision boundary suggests we may be capturing the heterogeneity of the two metastatic biopsies. These results support the presence of ER subtype heterogeneity as compared with orthogonal ctDNA clonality analysis and suggest that tumor subtype dynamics can be monitored during therapy.

## Discussion

In this study, we described the development of Griffin, a new framework and analysis tool for studying transcriptional regulation and tumor phenotypes. Griffin uses a novel cfDNA fragment length-specific normalization of GC-content biases that obscure chromatin accessibility information. We demonstrated that Griffin can be used to detect cancer from low pass WGS with high accuracy. We also developed an approach to perform ER subtyping in breast cancer from ULP-WGS, which to our knowledge is the first time that ER phenotype prediction has been shown from ctDNA.

Griffin is versatile and can be used for various applications in cancer. We highlighted cancer detection, tissue-of-origin, and tumor subtype use-cases. However, Griffin can also be used for any biological comparison where transcriptional regulation and chromatin accessibility differences can be delineated. The applications described here use TFBSs from chromatin immunoprecipitation sequencing (ChIP-seq) and accessible chromatin sites from ATAC-seq. However, Griffin differs from existing methods due to its ability to analyze custom sites of interest that are specific to any biological context. These sites may be obtained from external sources and different assays, such as ChIP-seq, DNase I hypersensitivity, ATAC-seq or cleavage under targets and release using nuclease (CUT&RUN). As additional epigenetic data are collected by the cancer research community, including from single-cell experiments^51,52^, Griffin will be integral for advancing tumor phenotype studies from liquid biopsies.

Griffin is optimized for the analysis of ULP-WGS (0.1x) of cfDNA, while other nucleosome profiling methods have focused on deeper coverage sequencing. Griffin takes advantage of analyzing the breadth of sites as opposed to individual loci, which was inspired by a similar strategy used by Ulz et al^42^. We show that Griffin has better performance for both detecting cancer and predicting ER status from ULP-WGS data when compared to the Ulz method, because of its novel bias correction and versatility to analyze any set of genomic regions. However, Griffin is not limited to low coverage data. Increased cfDNA sequencing coverage can allow for analysis of specific gene promoters and cis-regulatory elements and may be able to inform gene expression.^31^ While recent studies show the promise of cfDNA methylation and cfRNA analysis for tumor phenotype analysis and cancer detection,^53–59^ these analytes may be challenging to isolate from clinical specimens or require specialized assays. Griffin provides a cost-effective and scalable method requiring only standard low coverage WGS of cfDNA, which can be more rapidly incorporated into existing platforms to predict clinical cancer phenotypes.

A limitation of the binary ER classification (ER+ or ER-) is the decreased accuracy for samples with lower tumor fraction (0.05 to 0.1); however, patients with cfDNA tumor fraction ≥ 10% have poorer prognosis^60^ and would benefit more from tumor monitoring. It may be possible to improve performance of ER subtyping for lower tumor fraction samples with additional sequencing depth or joint analysis of multiple cfDNA timepoints from the same patient.

The application of Griffin to predict ER status from cfDNA of MBC patients led to interesting insights into tumor heterogeneity and potential explanations for misclassified predictions. Intriguingly, we noticed that for the patients with ER-tumors by IHC, ER+ predictions were significantly enriched when the primary tumor was ER+. Moreover, we observed that the predicted ER probability closely matched the clonal dynamics from somatic mutation in six patients. Two of these patients had a change in predicted ER status, potentially suggesting the presence of metastases of both subtypes. Importantly, while this subtype heterogeneity and switching would typically not be captured from a single metastatic biopsy, our results demonstrate the possibility of using ER probability to monitoring subtype heterogeneity over time during therapy using ctDNA.

We focus our breast cancer subtyping on ER prediction because its status has important utility in predicting likely benefit to endocrine therapy.^61^ While PR expression is also determined in the clinic and ER-/PR+ tumors are considered hormone receptor positive, these are rare, not reproducible or less useful for prognosis.^62^ In our cohort, only 2 of 139 (1.4%) patients were ER-/PR+. HER2 overexpression is important relevant for prognosis and determining treatment such as trastuzumab.^63^ However, we were unable to identify sufficient number of open chromatin sites that were specific for distinguishing HER2 status. Since ERBB2 (encodes the HER2 protein) is amplified in ∼20% breast cancers, one can instead assess ERBB2 copy number amplification from ctDNA genomic analysis.^64^ Alternatively, a model to predict PAM50 status could be useful as this may be a better indicator of prognosis than ER/PR/HER2 IHC alone.^65^

The Griffin framework is a unique advance on our previous method to analyze genomic alterations and estimate tumor fraction from ULP-WGS of cfDNA.^10^ Together, these methods form a suite of tools to establish a new paradigm to study both tumor genotype and phenotype from ULP-WGS of cfDNA. Griffin has the potential to reveal clinically relevant tumor phenotypes, which will support the study of therapeutic resistance, inform treatment decisions, and accelerate applications in cancer precision medicine.

## Methods

### Griffin: Site filtering

Prior to performing nucleosome profiling, we filtered all site lists by mappability to remove regions that had low or uneven coverage due to inability to map reads. We used mappability data from the hg38 Umap multi-read mappability track for 50bp reads downloaded from the UCSC genome browser^66^ (downloaded from here https://hgdownload.soe.ucsc.edu/gbdb/hg38/hoffmanMappability/k50.Umap.MultiTrackMappability.bw). To perform this filtration, we developed the ‘griffin_filter_sites’ pipeline. This pipeline takes a mappability file, a list of sites, a window to examine around each site, and a mappability threshold. We used a window of -5,000 to +5,000 bp around each site. Within this window, we calculated the mean mappability value using pyBigWig (https://github.com/deeptools/pyBigWig). We then excluded sites with a mean mappability below the threshold of 0.95 and retained highly mappable sites for further analysis.

### Griffin: GC bias calculation

GC content influences the efficiency of amplification and sequencing leading to different expected coverages (coverage bias) for fragments with different GC contents and fragment lengths. This is called GC bias and is unique to each sample. We calculated the GC bias of each bam file using a custom method similar to that demonstrated in Benjamini and Speed 2012^44^ and implemented in deepTools^67^. However, unlike this existing approach, which assumes that all fragments have the same length, our approach calculates a separate GC bias curve for every fragment length within a specified range. This is helpful for cfDNA where different samples may have different fragment size distributions. Prior to performing GC bias calculation, we identified all mappable, non-repetitive regions of the genome. We used pybedtools to find the mappable regions (defined as mappability score = 1) from the hg38 mappability track (described in the section on site filtering) and exclude the repetitive regions from the UCSC hg38 repeat masker track^68^ (downloaded from the UCSC table browser: http://genome.ucsc.edu/cgi-bin/hgTables). We then examined all mappable, non-repetitive regions of the genome and, for each fragment length, counted the number of times each GC content is observed in possible fragments overlapping those regions. These counts for each fragment length are the ‘genome GC frequencies’. We then developed the ‘griffin GC bias’ pipeline to compute the GC bias in a given bam file. The pipeline takes a bam file, bedGraph file of valid (mappable, non-repetitive) regions, and genome GC frequencies for those regions. For each given sample, we fetched all reads aligning to mappable, non-repetitive regions on autosomes using pysam (https://github.com/pysam-developers/pysam)^69^. We counted the number of observed reads for each length and GC content, excluding reads with low mapping quality (<20), duplicates, unpaired reads, and reads that failed quality control. These read counts are the ‘GC counts’ for that sample. We then divided the GC counts by the GC frequencies to obtain the GC bias for that bam file and normalized the mean GC bias for each fragment length to 1, resulting in a GC bias value for every combination of fragment size and GC content (except those that are not observed in the genome). We then smoothed the GC bias curves. For each fragment size we took all GC bias values for fragments of a similar length (+/- 10 bp). We sorted these values by the GC content of the fragment to create a vector of GC bias values for similar sized fragments. We then smoothed this vector by taking the median of k nearest neighbors (where k = 5% of the vector length or 50, whichever is greater) and repeated for each possible fragment length. We then normalized to a mean GC bias of 1 for each possible fragment length to generate a smoothed GC bias value for every possible fragment length and GC content observed in the genome.

### Griffin: Nucleosome profiling

We designed the griffin nucleosome profiling pipeline to perform nucleosome profiling around sites of interest. This pipeline takes a bam file and site list, and assorted other parameters described below. For a given bam file and site list, we fetched all reads in a window (−5000 to +5000bp) around each site using pysam (excluding those that failed quality control measures). We then filtered reads by fragment length and selected those in a range of fragment lengths (typically 100-200 bp unless otherwise specified). For each read, we determined the GC bias for each fragment and assigned a weight of 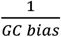 to that fragment and identified the location of the fragment midpoint. We split the site into 15bp bins and summed the weighted fragment midpoints in each bin to get a GC corrected midpoint coverage profile (see Fig. 1b for a schematic). We repeated this for every site on the site list and took the mean of all sites to generate the coverage profile for that site list. To make samples with different depths comparable, we normalized the coverage profile to a mean coverage of 1. We then smoothed the coverage profiles using a Savitzky-Golay filter with window length 165bp and polynomial order of 3.

### Griffin: Nucleosome profile feature quantification

To quantify coverage profiles, we extracted 3 features from each coverage profile. First, we calculated the ‘mean coverage’ value +/- 1000 bp from the site. Second, we calculated the coverage value at the site (+/- 30bp). And third, we calculated the amplitude of the nucleosome peaks surrounding the site by using a Fast Fourier Transform (as implemented in Numpy^70^) on the window +/-960 bp from the site and taking the amplitude of the 10^th^ frequency term. This window and frequency were chosen due to the observed nucleosome peak spacing at an active site (190bp) which results in approximately 10 peaks in the window +/-960bp.

### Early-stage cancer and healthy donor cfDNA samples

Whole genome sequencing (WGS) cfDNA from patients with various types of early stage cancer and healthy donors were obtained from an existing dataset published in Cristiano et al^38^. Bam files were downloaded from EGA (dataset ID: EGAD00001005339). This data consisted of 1-2x low pass whole genome sequencing from 100bp paired end Illumina sequencing reads. For our analyses, we used a subset of samples with 1-2X WGS of cfDNA from 208 cancer patients with no previous treatment and 215 healthy donors. These are the samples used for the cancer detection analysis in Cristiano et al. cfDNA tumor fraction was estimated using ichorCNA^10^. An hg38 panel of normal (PoN) with a 1mb bin size was created using all 260 healthy donors in the dataset. ichorCNA was then run on all cancer and healthy samples to estimate tumor fraction. ichorCNA_fracReadsInChrYForMale was set to 0.001. Defaults were used for all other settings.

### Metastatic breast cancer (MBC) and healthy donor cfDNA samples

WGS of cfDNA from patients with metastatic breast cancer (MBC) and healthy donors were obtained from an existing dataset published by Adalsteinsson and colleagues^10^. Bam files were downloaded from dbGaP (accession code: phs001417.v1.p1). This data consisted of ∼0.1x ultra-low pass whole genome sequencing (ULP-WGS) from 100bp paired end Illumina sequencing reads. For our analyses, we used a subset of 254 samples with >0.1X coverage WGS, >0.05X tumor fraction and known estrogen receptor (ER) status. Of these 254 samples 132 were ER positive (from 74 unique patients) and 122 were ER negative (from 65 unique patients). Coverage and tumor fraction metrics were obtained from the supplemental data in the publication^10^. Primary and metastatic ER and PR status determined by immunohistochemistry. Additionally, we used deep (9-25X) WGS from two MBC patients (MBC_315 and MBC_288) from the same source and deep (17-20X) WGS from two healthy donors (HD45 and HD46) from the same source for designing and demonstrating the pipeline.

For training and assessing the ER status classifier we labeled each sample as ER+ or ER-using information about the ER status from medical records. If metastatic ER status was known, the sample was labeled according to this status. If metastatic ER status was not known, the sample was labeled according to the primary tumor ER status (20 samples from 11 patients). ER low samples (9 samples from 5 patients) were labeled ER positive for the purpose of the binary classifier. For three patients (MBC_1405, MBC_1406, MBC_1408), we had information about multiple metastatic biopsies with different ER statuses. In these cases, we used the last biopsy taken for the purpose of the binary ER status classifier.

### Human Subjects

WGS of cfDNA samples from patients with MBC were obtained from an existing study as described above^10^. Additional information, including primary ER status, metastatic ER status, and survival time, was abstracted from the medical records. Use of this data was approved by an institutional review board (Dana-Farber Cancer Institute IRB protocol identifiers 05-246, 09-204, 12-431 [NCT01738438; Closure effective date 6/30/2014]).

### Sequence data processing

All sequencing data used in this study was realigned to the hg38 version of the human genome (downloaded from http://hgdownload.soe.ucsc.edu/goldenPath/hg38/bigZips/hg38.fa.gz). Bam files were unmapped from their previous alignment using Picard SamToFastq.^71^ They were then realigned to the human reference genome according to GATK best practices^72^ using the following procedure. Fastq files were realigned using BWA-MEM.^73^ Files were then sorted with samtools^74^, duplicates were marked with Picard, and base recalibration was performed with GATK, using known polymorphisms downloaded from the following locations: https://console.cloud.google.com/storage/browser/genomics-public-data/resources/broad/hg38/v0/Mills_and_1000G_gold_standard.indels.hg38.vcf.gz and https://ftp.ncbi.nih.gov/snp/organisms/human_9606_b151_GRCh38p7/VCF/GATK/All_20180418.vcf.gz.

### Transcription factor binding site (TFBS) selection

Transcription factor binding sites (TFBSs) were downloaded from the GTRD database^45^. This database contains a compilation of ChIP seq data from various sources. For our analyses, we used the meta clusters data (version 19.10, downloaded from https://gtrd.biouml.org/downloads/19.10/chip-seq/Homo%20sapiens_meta_clusters.interval.gz). This contains meta peaks observed in one or more ChIP seq experiments. The GTRD database contains some ChIP seq experiments for targets that are not transcription factors (TFs). These were excluded by comparing against a list of TFs with known binding sites in the CIS-BP database^75^ (v2.00 downloaded from http://cisbp.ccbr.utoronto.ca/bulk.php). TFBS were then filtered by mappability as described above (Griffin: Site Filtering). The site position was identified as the mean of ‘Start’ and ‘End’. TFs with less than 10,000 highly mappable sites on autosomes were excluded. For each remaining TF, the top 10,000 highly mappable sites were selected by choosing those with the highest ‘peak.count’ (number of times that peak has been observed across all experiments).

### DNase I hypersensitivity site selection

DNase I hypersensitivity sites for a variety of tissue types were downloaded from https://zenodo.org/record/3838751/files/DHS_Index_and_Vocabulary_hg38_WM20190703.txt.gz^76^. These sites were split by tissue type for a total of 16 site lists. They were filtered by mappability as described above (Griffin: Site Filtering) using the ‘summit’ column as the site position. The highly mappable sites were sorted by the number of samples where that site had been observed (‘numsamples’) and the top 10,000 most frequently observed sites were selected for each tissue type.

### ATAC-seq site selection for ER subtyping

Assay for transposase-accessible chromatin using sequencing (ATAC-seq) site accessibility for primary breast cancer samples from The Cancer Genome Atlas (TCGA) were downloaded from the TCGA ATAC-seq hub (https://atacseq.xenahubs.net/download/brca/brca_peak_Log2Counts_dedup)^47^. The locations of these sites and patient metadata were obtained from the supplemental tables in the paper^47^. These ATAC-seq sites were filtered for mappability as described above (Griffin: Site Filtering), using the mean of the Start and End columns as the peak position. High mappability sites on autosomes were kept for further analysis for a total of 142,490 sites. Differentially accessible sites between ER+ (n=44) and ER-(n=15) tumors were identified by using a Mann-Whitney U test. P values were corrected for multiple testing using the Benjamini/Hochberg procedure using statsmodels^77^ and sites with a q-value <0.05 were selected. Additionally, selected sites were further filtered based on the log2 fold change between ER+ and ER-tumors. Sites with a log2 fold change >0.5 were classified as ER+ specific, while sites with a log2 fold change <-0.5 were classified as ER-specific. These site lists were further split into sites shared with hematopoietic cells and those not shared with hematopoietic cells. Hematopoietic sites were obtained from a database of single cell ATAC-seq data^48^ (GEO accession number: GSE129785, peak file available here: https://ftp.ncbi.nlm.nih.gov/geo/series/GSE129nnn/GSE129785/suppl/GSE129785%5FscATAC%2DHematopoiesis%2DAll%2Epeaks%2Etxt%2Egz). Hematopoietic peaks were lifted over to hg38 using the UCSC liftover command line tool and sites that changed size during liftover (0.2% of peaks) were discarded. BRCA ATAC-seq sites that overlapped with Hematopoietic sites (Overlapping peaks were defined as site centers being within 500bp of one another) this was performed using pybedtools intersect^78,79^. This resulted in a total of 4 differential site lists: ER positive sites that were not shared with hematopoietic cells (15,142 sites), ER positive sites that were shared with hematopoietic cells (12,217 sites), ER negative sites that were not shared with hematopoietic cells (12,151 sites), and ER negative sites that were shared with hematopoietic cells (12,710 sites).

We then overlapped these differential ATAC-seq site lists with the top 10,000 sites for each of 338 transcription factors (TFs) using pybedtools intersect. An overlapping pair of sites was defined as having <500bp between site centers. Each differential ATAC-seq site list was compared against each list of TFBSs and the total number of ATAC sites overlapping one or more TFBS on the given list was recorded.

### Assessment of Griffin before and after GC correction

#### Tumor fraction correlations at TFBS

For 191 MBC ULP samples with >0.1 tumor fraction, nucleosome profiling with and without GC correction was performed on the top 10,000 sites for each of 338 transcription factors (TFs). For each TF, the relationship between central coverage and tumor fraction was modeled using scipy.stats.linregress^80^ producing a Pearson correlation (r) and line of best fit. Root mean squared error (RMSE) was calculated from the line of best fit. This was performed both before and after GC correction as illustrated for Lyl-1 in Fig. 2e. For all 338 TFs, the RMSE values before and after GC correction were compared using a Wilcoxon signed-rank test (two-sided).

#### Mean absolute deviation (MAD) at TFBS

For 215 healthy donors, nucleosome profiling with and without GC correction was performed on the top 10,000 sites for each of 338 TFs. For each TF, the MAD of the central coverage values was calculated both before and after GC correction. For all 338 TFs, the MAD values before and after GC correction were compared using a Wilcoxon signed-rank test (two-sided).

### Machine learning, bootstrapping, and performance evaluation procedure

To detect cancer, predict tissue type, or predict ER subtype, we used logistic regression with Ridge regularization (i.e. L2 norm) as implemented in scikit-learn^81^. All feature values were scaled to a mean of 0 and a standard deviation of 1 prior to performing bootstrapping and fitting the models. We used the following bootstrapping procedure to train and assess the performance of our models. First, we selected n samples with replacement from the full set of n samples and used this as a training set. Samples that weren’t selected were used as the test set. We then used 10-fold cross-validation on the training set to select the parameter ‘C’ (inverse of the regularization strength) from the following options: 10^−5^, 10^−4^, 10^−3^, 10^−2^, 10^−1^,10^0^, 10^1^, 10^2^. To account for class imbalances in the data we used set the ‘class weight’ parameter to ‘balanced’ to adjust the sample weighs inversely proportional to the class frequencies. We trained a final model on all the training data using the selected regularization strength. Finally, we tested this model on the test set and recorded the performance (accuracy and AUC values) and probabilities from each sample. Then, a new training set was selected, and the procedure was repeated for 2000 iterations (for cancer detection and tissue of origin analysis) or 1000 iterations (for breast cancer subtyping). After completing the bootstrap iterations, we calculated the AUC and accuracy from each bootstrap iteration and used these to generate the mean and 95% confidence interval around each of these values. To visualize the mean ROC curve, we used the median probability from all bootstraps where that sample was included in the test set. For further downstream analyses, we used this same median probability.

### Features used for cancer detection classification

To detect cancer, we applied the logistic regression approach described above and built four different models using four different sets of features extracted from the pan cancer patient samples and healthy donor samples. First, we performed nucleosome profiling in these samples (selecting fragments 100-200bp in length) on the 338 selected TFs from the GTRD database. We extracted three features (as described above) from each coverage profile for a total of 1,014 features.

Second, we performed nucleosome profiling on these same samples and sites but selected only ‘short’ fragments (35-150bp) to be counted in the nucleosome profiles.

Third, we downsampled these samples to ∼0.1x coverage (procedure described below) and performed nucleosome profiling for the same 338 TFs selecting fragments 100-200bp in length. Fourth, we used the original (not downsampled) samples and performed nucleosome profiling at the 16 tissue-specific DHS site lists described above. We extracted the same 3 features from each site profile for a total of 48 features.

### Tissue of origin prediction

For tissue of origin prediction, we used the nucleosome profiles from the 338 TFs in the 1-2X coverage (not downsampled) cancer samples using 100-200bp fragments. We excluded 1 duodenal cancer sample as this was the only sample from that cancer type. This left us with 207 cancer samples from 7 different cancer types: bile duct (n= 25), breast (n=54), colorectal (n=27), gastric (n=27), lung (n=12), ovarian (n=28), and pancreatic (n=34). We built a multinomial logistic regression model to predict the cancer tissue of origin for each sample using the same bootstrapping strategy described above. We ran this for 2000 iterations. For each iteration, we calculated the accuracy of the top prediction as well as the top two predictions.

### Downsampling of pan-cancer and healthy donor cfDNA sequencing data

1-2x WGS of pan-cancer patient and healthy donor bam files aligned to hg38 were downsampled using Picard DownSampleSam. The probability used by DownSampleSam was calculated based on a target of 2,463,109 read pairs which resulted in approximately 0.11x coverage as calculated by Picard CollectWgsMetrics. Downsampled bam files were realigned to hg19 for use in the Ulz pipeline. The realignment procedure was the same as above but using the hg19 genome (downloaded from https://hgdownload.soe.ucsc.edu/goldenPath/hg19/bigZips/hg19.fa.gz) and hg19 known polymorphic sites for base recalibration (downloaded from ftp://gsapubftp-anonymous@ftp.broadinstitute.org/bundle/hg37/Mills_and_1000G_gold_standard.indels.hg37.vcf.gz and ftp://ftp.ncbi.nih.gov/snp/organisms/human_9606_b151_GRCh37p13/VCF/GATK/All_20180423.vcf.gz).

### ER status classification in the MBC cohort

To predict ER status, we applied the logistic regression approach described above to features extracted from the MBC patient samples. Because some patients had multiple samples, we modified the bootstrapping procedure to select 139 patients (rather than samples) with replacement from a full set of 139 patients. For each selected patient, all samples from that patient were added to the training set (If a patient was selected multiple times, all their samples were included multiple times). This ensured that separate samples from the same patient (biological replicates) could not appear in both the training and test set. Samples from patients that weren’t selected were used as the test set.

Using these training and tests sets, we built three different models based on three different sets of features. First, we applied nucleosome profiling using 100-200bp fragments to the 338 TFs from GTRD and extracted 3 features per profile for a total of 1014 features. Second, we applied nucleosome profiling using 100-200bp fragments to the 4 ER differential ATAC seq lists and extracted 3 features per profile for a total of 12 features. Lastly, we applied nucleosome profiling using 35-150bp fragments to the 4 ER differential ATAC seq lists and extracted 3 features per list for a total of 12 features.

For evaluating the models, we only included the first timepoint for each patient in the test set when calculating the accuracy and AUC for each bootstrap iteration. This prevented a small number of patients with many samples from having a large impact on the scores.

### ER probability comparison between patients with and without ER loss using analysis of covariance (ANCOVA)

To determine whether the probability of ER+ for the patients with ER loss (primary ER+, metastatic ER-) were significantly different from the probability of ER+ for the patients with ER-primary and metastasis disease, we performed an analysis of covariance (ANCOVA) as implemented in Pingouin^82^. Probability of ER+ was the dependent variable, primary tumor status was the independent variable (‘between’), and tumor fraction was a covariate. While we found that tumor fraction was significantly related to the ER probability (p=0.03, F= 5.02, degrees of freedom = 1), we also found a significant difference (p=0.014, F = 6.48, degrees of freedom = 1) between the ER loss and ER-unchanged patients.

### Transcription factor profiling using pipeline from Ulz et al

We downloaded the Transcription Factor Profiling pipeline published by Ulz and colleagues from Github (https://github.com/PeterUlz/TranscriptionFactorProfiling)^42^ and ran it using the following procedure as described in the paper. hg19 aligned bam files were used because the pipeline was written to for this version of the genome. Scripts were modified so that they worked in python3. We trimmed the reads in each bam to 60bp using ‘trim from bam single end’ with modifications to skip unaligned reads. We ran ichorCNA on the original (untrimmed) bam using the default ichorCNA settings for hg19 except the bin size, which was modified to 50,000bp and no panel of normals. We then ran the transcription factor profiling analysis on the trimmed bam using the script run_tf_analyses_from_bam.py with options ‘-calccov’ and ‘-a tf_gtrd_1000sites’ and the ichorCNA corrected depth file as the ‘-norm-file’. This ran transcription factor profiling on 1,000 sites for each of 504 TFs. Finally, we ran the scoring pipeline. We used the high frequency amplitude (‘HighFreqRange’) for each of the 504 TFs in the accessibility output file (Accessibility1KSitesAdjusted.txt) as the features for a logistic regression model using the same bootstrapping scheme described above.

### Clonality analysis

For 6 patients with high tumor fractions, multiple samples, and triple negative breast cancer, data on clonal dynamics in the ctDNA was available from a previous study^49^ (results downloaded from: https://gitlab.com/Zt_Weber/narrow-interval-clonal-structure-mbc/-/tree/master/PyClone-Multisample-Final/pyclone_output_tables). In the study, somatic alterations were identified from both WES and targeted panel sequencing using GATK-Mutect2. Using these alterations, clonal dynamics were modeled using the PyClone^50^ package. The cellular prevalence estimate represents the proportion of the sample that contains somatic mutation. PyClone reports clusters of somatic mutations; cellular prevalence of these clusters is shown in the results.

## Data Availability

Sequencing data used in this study was obtained from dbGaP (accession phs001417.v1.p1) and EGA (dataset ID EGAD00001005339).

## Code availability

Griffin software and the subtype classifier tool can be obtained from https://github.com/adoebley/Griffin. Code for analysis and machine learning models can be accessed at https://github.com/adoebley/Griffin_analyses.

## Acknowledgments

We thank the many patients and their families for their generosity in contributing to this study. We also thank Patricia Galipeau, Anat Zimmer and the Ha laboratory for helpful discussion and critical reading of the manuscript. This work was supported by the National Institute of Health K22 CA237746 (to G.H.), the V Foundation Scholar Grant (to G.H.), Prostate Cancer Foundation Young Investigator Award (to G.H.), the Fund for Innovation in Cancer Informatics Major Grant (to G.H.). This research was also supported by the NIH/NCI Cancer Center Support Grant P30 CA015704, Brotman Baty Institute for Precision Medicine, NIH (P50 CA097186; R01 CA2344715 to P.S.N; K08 CA252649 to H.A.P.; P50 CA168504 to H.A.P.; K12 CA076930 to J.H.; T32 HL007093 to J.H.), CDMRP W81XWH-18-10406 (to P.S.N), Komen Breast Cancer Foundation Catalyst Research Grant (to H.A.P.). Scientific Computing Infrastructure was funded by ORIP Grant S10OD028685.

## Author contributions

A-L.D. and G.H. conceived the study, designed all the experiments, and wrote the manuscript. A-L.D. developed and implemented the Griffin method and performed all the analysis. M.K., H.L., A.E.C, C.K., A.C.H.H., K.C. contributed to the analysis. K.S., H.A.P, D.G.S. provided clinical data. Z.T.W. provided clonality results. J.H., R.D.P., N.D.S., M.A., J.R. contributed to analysis discussions. P.P., V.A.A., P.S.N., H.A.P., D.G.S., D.M. contributed to discussions, provided guidance and interpretation of results. G.H. supervised the study. All authors reviewed and edited the manuscript.

